# Modulation of binocular rivalry with rapid monocular visual stimulation

**DOI:** 10.1101/2020.05.28.20116392

**Authors:** Dania Abuleil, Daphne McCulloch, Heidi Patterson, Benjamin Thompson

**Affiliations:** Department of Optometry and Vision Science, University of Waterloo, Waterloo, ON, N2L 3G1

**Author notes:** Corresponding Author: Dania Abuleil School of Optometry and Vision Science, University of Waterloo 200 Columbia St. W, Waterloo Ontario, N2L3G1.

**Keywords:** long-term potentiation (LTP), binocular rivalry, ocular dominance, visual tetanus, neuroplasticity

## Abstract

Rapid visual stimulation can increase synaptic efficacy by repeated synaptic activation. This long-term potentiation-like (LTP-like) effect can induce increase human visual cortex excitability. To examine the effect of rapid visual stimulation on perception, we tested the hypothesis that rapid monocular visual stimulation would increase the dominance of the stimulated eye in a binocular rivalry task. Participants (n = 25) viewed orthogonal 0.5 cpd gratings presented in a dichoptic anaglyph to induce binocular rivalry. Rivalry dynamics (alternation rate, dominance, and piecemeal durations) were recorded before and after 2 min of rapid monocular stimulation (9Hz flicker of one grating) or a binocular control condition (9Hz alternation of the orthogonal gratings viewed binocularly). Rapid monocular stimulation did not affect alternation rates or piecemeal percept duration. However, unexpectedly, rivalry dominance of the stimulated eye was significantly reduced. A control experiment revealed that this effect could not be explained by monocular adaptation. Together, the results suggest that rapid monocular stimulation boosts dominance in the non-stimulated eye, possibly by activating homeostatic interocular gain control mechanisms.

## Introduction

Long-term potentiation (LTP) is the process of strengthening synaptic efficacy through repeated activation. This fundamental mechanism of neuroplasticity involves a cascade of cellular and molecular changes and underpins the processes of learning and memory formation (Bliss & Lømo, 1973; Bliss, T.V.P. & Collingridge, 1993). Early research revealed that rapid electrical stimulation of presynaptic cells within the rabbit hippocampus induced a lasting increase in the response amplitude of postsynaptic cells (Bliss & Lømo, 1973). Subsequent studies demonstrated similar effects (Bröcher *et al*., 1992) and characterized the neurochemical changes that occurred as a result of the stimulation (Teyler & DiScenna, 1987; Hayashi *et al*., 2000). These changes included a rise in postsynaptic calcium, the release of glutamate, and the activation of N-methyl-D-aspartate (NMDA) receptors (Malenka & Nicoll, 1999). While LTP is typically induced using electrical stimulation *in vitro*, similar effects (a strengthening of neural responses following stimulation) have been reported in visual cortex using rapid visual stimulation in adult rats (Heynen & Bear, 2001; Frenkel *et al*., 2006) and in humans (Clapp *et al*., 2005; Teyler *et al*., 2005; Normann *et al*., 2007).

In human adults, 2-minutes of rapid visual stimulation of a high-contrast checkerboard increases the amplitude of the N1b component of visual evoked potentials (VEPs) (Teyler *et al*., 2005; Normann *et al*., 2007; Sanders *et al*., 2018). Rapid visual stimulation, sometimes referred to as visual tetanus, has been delivered in a number of ways including 9Hz flicker of checkerboard or grating stimuli and 2 Hz pattern reversal of checkerboard stimuli (Teyler *et al*., 2005; Normann *et al*., 2007). To account for the effect of visual adaptation that can reduce visual cortex excitability and VEP amplitude (Blakemore & Campbell, 1969), most studies of rapid visual stimulation include a period of eye closure that at least matches the duration of rapid visual stimulation (Magnussen & Greenlee, 1985). The effect of rapid visual stimulation on VEP amplitude is stimulus specific (Vassilev *et al*., 1994; Ross *et al*., 2008), reliant on NMDA receptors in animal models (Clapp *et al*., 2006), and may also involve an increase in glutamate receptor expression (Eckert *et al*., 2013), suggesting that it involves an LTP-like mechanism.

The majority of studies into rapid visual stimulation in humans have used electrophysiology or neuroimaging to measure visual cortex excitability before and after stimulation (Sanders *et al*., 2018). Therefore, the perceptual effects of rapid visual stimulation, if any, are not well understood. This is an important issue. If the LTP-like changes in cortical excitability induced by rapid visual stimulation can modulate perception, rapid visual stimulation may have therapeutic applications. For example, repetitive transcranial magnetic stimulation of the visual cortex can transiently improve visual functions such as contrast sensitivity in adults with amblyopia, a neurodevelopmental disorder of vision (Thompson *et al*., 2008; Clavagnier *et al*., 2013; Tuna *et al*., 2020). Like rapid visual stimulation, the effects of repetitive transcranial magnetic stimulation on cortical excitability likely involve LTP-like mechanisms (Hoogendam *et al*., 2010). Therefore, rapid visual stimulation may have similar effects and, unlike repetitive transcranial magnetic stimulation, can be delivered to the thalamocortical inputs from just one eye. This property may make repetitive visual stimulation particularly well suited for the treatment of amblyopia, which is characterised by a large imbalance in neural response between the two eyes (Barnes *et al*., 2001).

Two preliminary studies have reported behavioural effects of rapid visual stimulation. Beste et al. observed improved luminance discrimination following 40 minutes of 20 Hz rapid visual stimulation, whereas Clapp et al. observed a reaction time improvement, but no change in response accuracy, during a checkerboard detection task following 2 minutes of 9Hz stimulation (Beste *et al*., 2011; Clapp *et al*., 2012). In this experiment we further explore the behavioural effects of rapid visual stimulation by investigating the effect of monocular rapid visual stimulation on binocular rivalry.

Binocular rivalry is a form of bistable perception wherein conflicting monocular images stochastically compete for dominance when viewed dichoptically. The resulting percept can involve periods of complete perceptual dominance by one eye, and periods of a mixed, piecemeal percept whereby each eye dominates in different regions of the visual field (Wilson *et al*., 2001). In individuals with normal binocular vision, the periods of perceptual dominance are relatively equal between the two eyes. However, the relative dominance of each eye during binocular rivalry can be modulated by varying stimulus features such as size (Kang, 2009), colour (Stalmeier & de Weert, 1988), luminance (Hong & Shevell, 2008), orientation (Holmes *et al*., 2006) and spatial frequency (Fahle, 1982) between the two eyes.

In this study, we induced binocular rivalry by dichoptically presenting orthogonal, sinusoidal gratings. Dichoptic presentation was achieved using red/green anaglyphs. The aim of our first experiment was to determine suitable grating parameters. Specifically, we aimed to identify a stimulus configuration that generated minimal time spent in piecemeal and stable alternation rates across trials. In our second experiment we used this stimulus to assess whether monocular rapid visual stimulation modulates binocular rivalry dynamics and/or dominance durations in individuals with normal binocular vision. Our hypothesis was that rapid monocular visual stimulation would strengthen the cortical response to inputs from the stimulated eye and that this would increase the relative time spent perceiving the stimulus presented to the stimulated eye during binocular rivalry (i.e. increase the perceptual dominance of the stimulated eye). In a third experiment, we measured binocular rivalry before and after viewing a monocular static grating as a test of monocular visual adaptation.

## Materials and Methods

Three experiments were performed. Experiment 1 was designed to determine the parameters for the binocular rivalry stimulus. Experiment 2 was designed to determine the effect of rapid monocular stimulation on binocular rivalry dynamics. A subset of participants from Experiment

2 completed a third experiment to determine whether adaptation could explain the results of Experiment 2.

## Experiment 1: Stimulation parameters for binocular rivalry

### Participants

Nine adults (age range: 21–28 years) with self-reported normal binocular vision participated in a 1-hour binocular rivalry experiment. All participants were informed of the nature of the study before participation and provided written informed consent. The project was approved by the University of Waterloo Research Ethics Committee.

### Stimuli and Protocol

Orthogonally oriented sinusoidally modulated gratings were presented dichoptically (57cm viewing distance) within a circular field subtending 6.1 degrees of visual angle on a gamma corrected 24² Asus® 3D monitor. Dichoptic presentation was achieved using red/green anaglyph glasses. The space average luminance levels of the gratings were matched using a Chroma Meter CS-100^Ò^ photometer through the anaglyphic filters. Using a computer keyboard, participants continuously reported whether they perceived the grating presented to the left eye (left eye dominant), the grating presented to the right eye (right eye dominant), or a piecemeal percept of both gratings. Specifically, a keyboard key was allocated to each percept. Participants held down the key corresponding to their current percept and switched keys when their percept changed. The total duration of each percept as well as the number of alternations (a change from one percept to another) were analysed.

Participants completed 40 х 60 sec randomly sequenced trials – 5 trials for each combination of two grating orientation pairs (45/135° vs. 90/180°) and 4 spatial frequencies (0.5, 1, 1.5 or 2 cycles per degree); the spatial frequency of the gratings presented to each eye within a trial was always identical.

### Analysis

Binocular rivalry alternation rates were calculated for each trial separately by dividing the number of alternations (defined as any change in percept) by the total presentation time. Alternation rate calculations included piecemeal percepts. Alternation rates across all five trials were then averaged for each set of stimulus parameters. The cumulative duration of piecemeal percepts was also analysed. Ocular dominance indices were calculated for each participant as (time spent viewing with right eye - time spent viewing with left eye)/(time spent viewing with right eye + time spent viewing with left eye). To investigate the effect of spatial frequency and orientation on ocular dominance, the absolute ocular dominance values were analysed.

Data were tested for normality using the Shapiro-Wilkes paired-samples assumption test. Normally distributed data were analysed using repeated measures ANOVA and post-hoc paired t-tests. Skewed data were analysed using the Durbin test and post-hoc Wilcoxon signed-rank tests. Repeated measures ANOVAs or Durbin tests with factors of orientation (90/180 vs. 45/135) and spatial frequency (0.5 vs. 1.0 vs. 1.5 vs. 2.0 cpd) were conducted separately for alternation rate, piecemeal duration, and the absolute ocular dominance index. To determine whether stimulus orientation or spatial frequency affected the stability of binocular rivalry dynamics across trials, each participant’s standard deviation across trials for each combination of orientation and spatial frequency was calculated for alternation rate. Repeated measures ANOVAs with factors of orientation and spatial frequency were conducted on the standard deviation data.

## Experiment 2: Binocular rivalry following rapid monocular stimulation

### Participants

Twenty-five adults (mean age 25, range 19–33) with normal binocular vision based on stereopsis of ≤ 40 arc sec (The Fly Stereo Acuity Test® Vision Assessment Corporation) and normal or corrected-to-normal vision (0.1 logMAR or better in each eye) participated in the rapid monocular stimulation experiment. Exclusion criteria included any neurological condition or the use of psychoactive drugs. All participants were informed of the nature of the study before participation and provided written informed consent. The project was approved by the University of Waterloo Research Ethics Committee.

### Rivalry stimulus

The stimulus spatial frequency and orientation pair determined in experiment 1 (0.5cpd, 45/135°) was chosen for this experiment. Viewing conditions and the method of reporting binocular rivalry percepts were identical to experiment 1. Three 60-second trials of binocular rivalry were recorded before and after rapid monocular stimulation.

### Study design

We used a modified version of the rapid monocular stimulation protocol described by Teyler and colleagues (2005) (Figure 1). Within a repeated measures design, participants completed two study conditions on separate days: a rapid monocular visual stimulation condition, and a binocular control condition. Upon the first visit, participants completed either the rapid monocular stimulation condition or the binocular control condition, assigned randomly. Rapid monocular stimulation involved monocular viewing of only one of the two gratings that made up the binocular rivalry stimulus flickering on and off (50% duty cycle, on: high contrast grating on a luminance-matched grey surround; off: uniform grey field) at 9Hz for 2 minutes. The stimulated eye (left or right) was randomly selected for each participant and participants wore red/green glasses during the rapid monocular stimulation. The binocular control condition was identical except that the two gratings that made up the binocular rivalry stimulus were alternated in the center of the monitor at 9 Hz and viewed binocularly (no red/green glasses). In both the rapid monocular stimulation and binocular control conditions, the two minutes of visual stimulation was followed by two minutes of eye closure to minimize adaptation effects. Binocular rivalry measures were recorded before stimulation (pre) and after eyelid closure (post).

**Figure 1:**
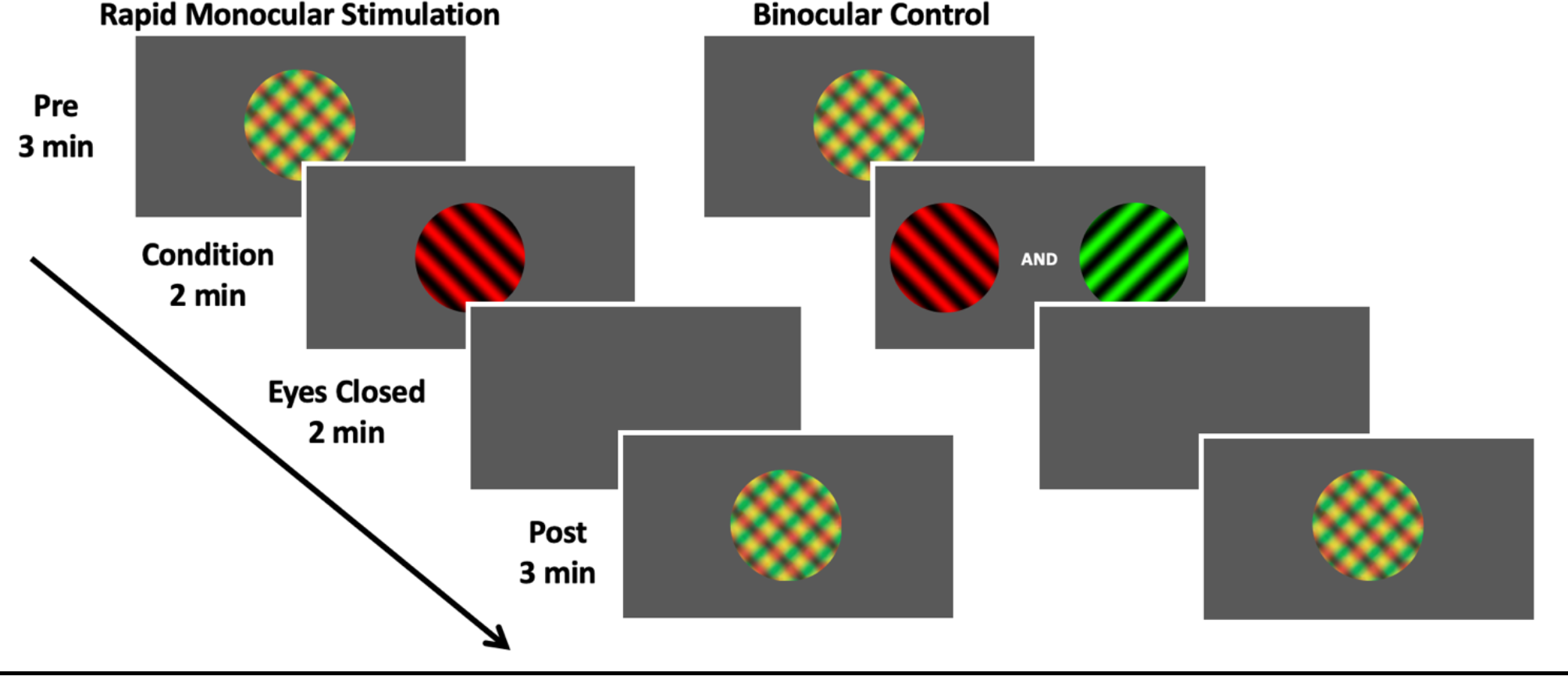
Schematic representation of Experiment 2 protocol. Plaid stimuli indicate binocular rivalry testing. In the rapid monocular stimulation condition, one the of the gratings that made up the plaid was presented monocularly and flickered at 9Hz. The stimulated eye (and therefore the red or green colour of the grating) was randomised. In this figure, the red grating is shown as an example. In the control binocular condition, the two gratings that made up the binocular rivalry stimulus were alternated at 9 Hz at the center of the screen and viewed binocularly.

### Analysis

The binocular rivalry measures were alternation rate, time spent in piecemeal, and ocular dominance index (all calculated as in experiment 1). Alternation rates and time spent in piecemeal across all three trials were averaged for each condition. An ocular dominance index was calculated for each participant based only on the duration of left eye dominant and right eye dominant percepts. Piecemeal percepts were not included in this analysis. In the rapid monocular stimulation condition this index was defined as: stimulated eye dominance duration - non-stimulated eye dominance duration)/(stimulated eye dominance duration + non-stimulated eye dominance duration); in the binocular control condition the ratio was calculated in the same way based on the eye stimulated in the monocular condition.

Data were analysed using parametric or non-parametric tests depending on normality as in experiment 1. ANOVAs or Durbin tests with factors of Condition (rapid monocular stimulation vs. control) and Time (pre vs. post stimulation) were conducted separately for alternation rate, piecemeal duration, and ocular dominance indices. Post-hoc testing was conducted using paired t-tests or the Wilcoxon signed-rank test.

## Experiment 3: Binocular rivalry following monocular adaptation

### Participants and methods

A subset of participants that completed experiment 2 who consented to and were available for additional testing (N = 12) completed experiment 3 on a separate day several months after completing experiment 2. Experiment 3 was a post-hoc experiment designed to investigate whether monocular adaptation could explain the results of experiment 2. The pre and post measurements of binocular rivalry used in experiment 3 were identical to those used in experiment 2. The monocular adaptation between these tests was a static monocular presentation of one of the gratings that made up the binocular rivalry stimulus for 2 minutes. The static grating was static and was presented to the same eye (left or right) that had been exposed to rapid monocular stimulation in experiment 2.

### Analysis

Two analyses were conducted. First, the results from the rapid monocular stimulation and control conditions in experiment 2 were reanalysed using only data from the subset of participants who completed experiment 3 to test whether the main finding from experiment 2 (reduced ocular dominance index for the stimulated eye in the rapid monocular stimulation condition but not the control condition) was present in the smaller sample. Wilcoxon signed-rank tests were used to compare the ocular dominance indices pre vs. post stimulation in the rapid monocular stimulation and control conditions. Second, a Wilcoxon signed-rank test was conducted on the data collected in experiment 3 to compare ocular dominance indices pre vs. post static visual adaptation of one eye.

## Results

### Experiment 1

For alternation rates, a repeated measures ANOVA showed no significant effects of Grating Orientation (p > 0.05; Figure 2A). However, a main effect of Grating Spatial Frequency was observed (F_1,8_ = 4.194, p = 0.027; Figure 2B, Table 1). Alternation rates were slowest at 0.5 cpd. Alternation rates for the 0.5 cpd stimulus differed significantly from the 1 cpd (t_8_ = −3.617, p =0.007) and 1.5 cpd (t_8_ = −3.485, p = 0.008) stimuli, but not the 2 cpd stimulus (t_8_ = −1.597, p =0.149). No significant effects of Grating Orientation or Grating Spatial Frequency were observed for piecemeal duration or for the standard deviations of alternation rate (all F < 3.903, all p >0.069). Absolute values of ocular dominance indices were not normally distributed. As a result, the Durbin test was conducted and showed no significant effect of Grating Orientation (F_1_ =0.130, W = −567.8, p = 0.716) or Grating Spatial Frequency (F_1_ = 2.641, W = −21.9, p = 0.062) on ocular dominance index. Based on these results, a spatial frequency of 0.5 cpd was chosen for experiment 2 because this spatial frequency induced the slowest alternation rates. The oblique orientations (45/135) were chosen for experiment 2 arbitrarily.

**Figure 2:**
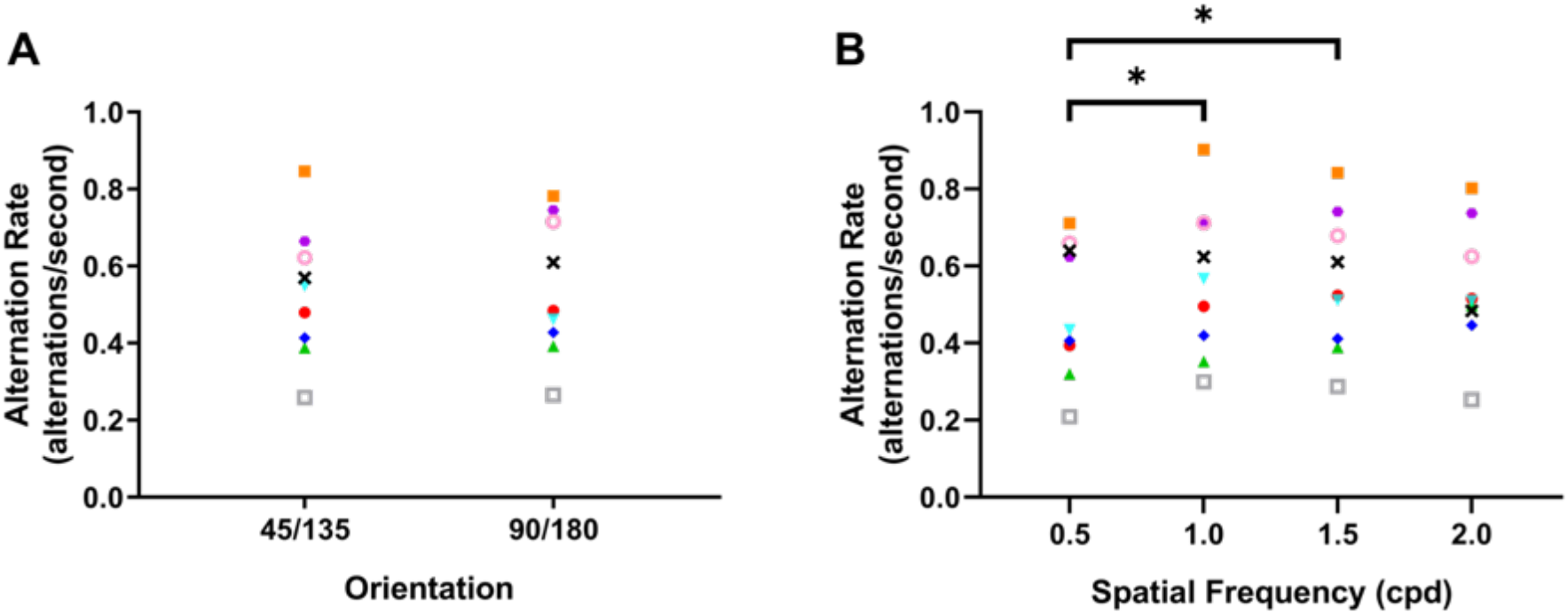
Rivalry alternation rates for experiment 1. (A) Orientation with the mean alternation rates for each individual participant collapsed across spatial frequency. (B) Spatial frequency with mean alternation rates for each individual participant collapsed across orientation. Each color signifies a different participant (n = 9). (*) indicates significant differences for post hoc paired t-tests p< 0.05.

**Table 1:** Experiment 1: Effects of stimulus parameters on binocular rivalry.

| Spatial Frequency | Alternation rate*<br>(Hz) | Piecemeal*<br>(time/60s) | Absolute Ocular<br>Dominance Index* |
| --- | --- | --- | --- |
| 0.5 cpd | 0.49 ($\pm 0.18$) | 0.06 ($\pm 0.02$) | 0.11 ($\pm 0.07$) |
| 1.0 cpd | 0.57 ($\pm 0.19$) | 0.10 ($\pm 0.03$) | 0.11 ($\pm 0.04$) |
| 1.5 cpd | 0.56 ($\pm 0.18$) | 0.12 ($\pm 0.04$) | 0.16 ($\pm 0.10$) |
| 2.0 cpd | 0.54 ($\pm 0.16$) | 0.13 ($\pm 0.04$) | 0.20 ($\pm 0.16$) |

\* grand mean between subjects followed by the mean of the within subjects' standard deviations.

### Experiment 2

Neither alternation rates nor ocular dominance indices were normally distributed. Therefore, nonparametric statistics were adopted. The median values ± interquartile range pre and post rapid monocular stimulation were 0.60 ± 0.24 Hz and 0.56 ± 0.24 Hz for alternation rates, 8.46 ± 10.13 s and 11.96 ± 12.33 s for time spent in piecemeal, and 0.02 ± 0.12 and −0.05 ± 0.08 for ocular dominance indices (Figure 3). For the binocular control condition, medians pre and post stimulation were 0.65 ± 0.28 and 0.61 ± 0.27 for alternation rates, 12.51 ± 11.71 s and 13.71 ±13.95 s for time spent in piecemeal, and −0.01 ± 0.09 and −0.02 ± 0.16 for ocular dominance indices. Rapid monocular stimulation did not alter binocular rivalry alternation rates (Durbin test: no effect of Condition [rapid monocular stimulation vs. binocular control]; F_1_ = 3.137, W = −17.9, p = 0.081), or the duration of piecemeal percepts (Durbin test: no effect of Condition [rapid monocular stimulation vs. binocular control]; F_1_ = 3.229, W = −18.1, p = 0.077). However, rapid monocular stimulation shifted the ocular dominance index in favour of the non-stimulated eye (Durbin test: significant effect of Condition [rapid monocular stimulation vs. binocular control]; F_1_ = 5.332, W = −18.8, p = 0.025). Post hoc Wilcoxon signed-rank tests revealed that the effect was associated with a significant shift in ocular dominance index towards the non-stimulated percept for the rapid monocular stimulation condition (W = 248.0, p = 0.005; Figure 4). In other words, rapid monocular stimulation decreased the time spent viewing the percept for the stimulated eye relative to that for the non-stimulated eye. There was no change in ocular dominance index for the binocular control condition (W = 134.5, p = 0.668).

**Figure 3:**
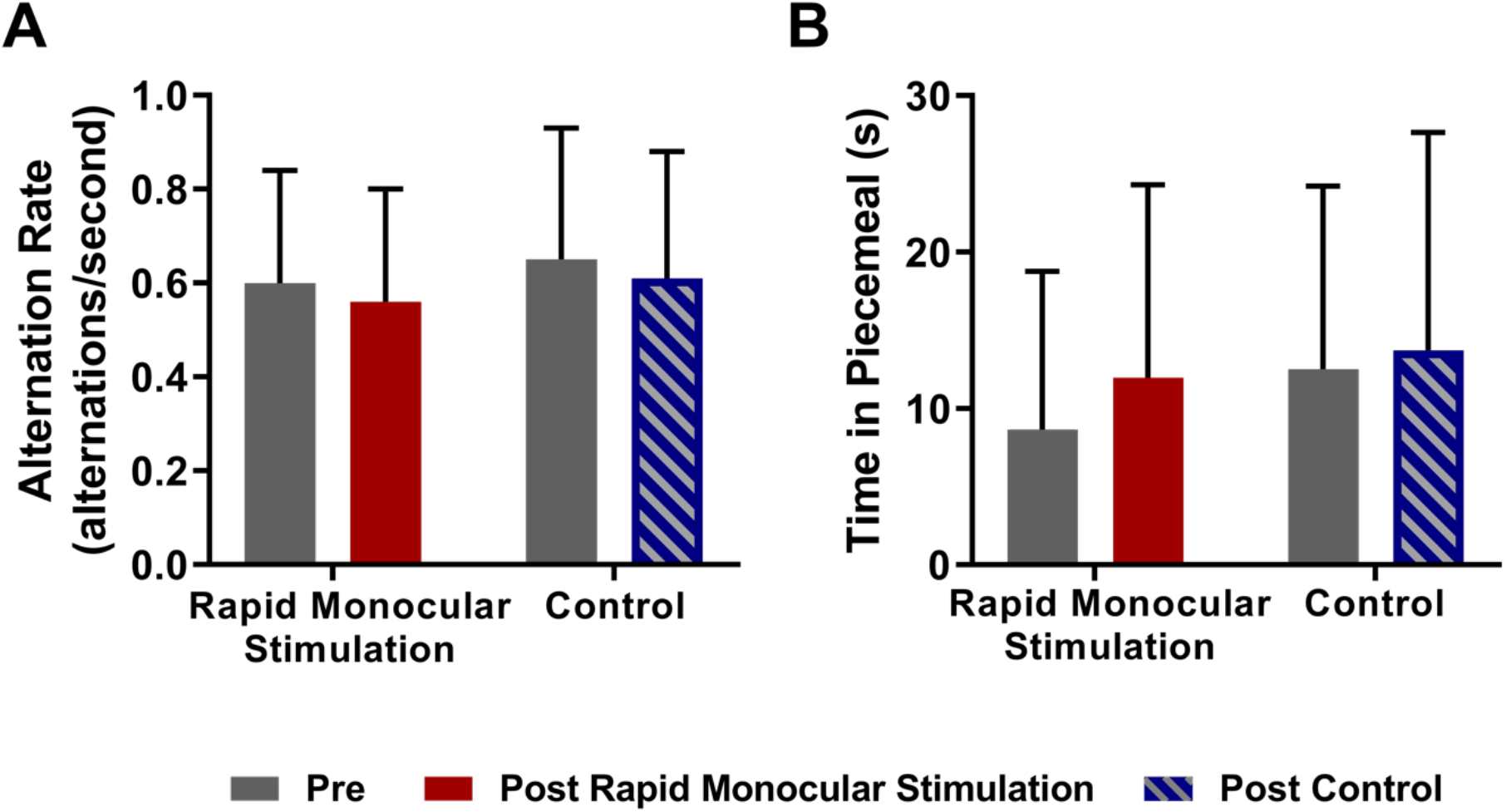
Median alternation rates (A) and time spent perceiving piecemeal (B) for the rapid monocular stimulation and binocular control conditions in experiment 2. Error bars = IQR.

**Figure 4:**
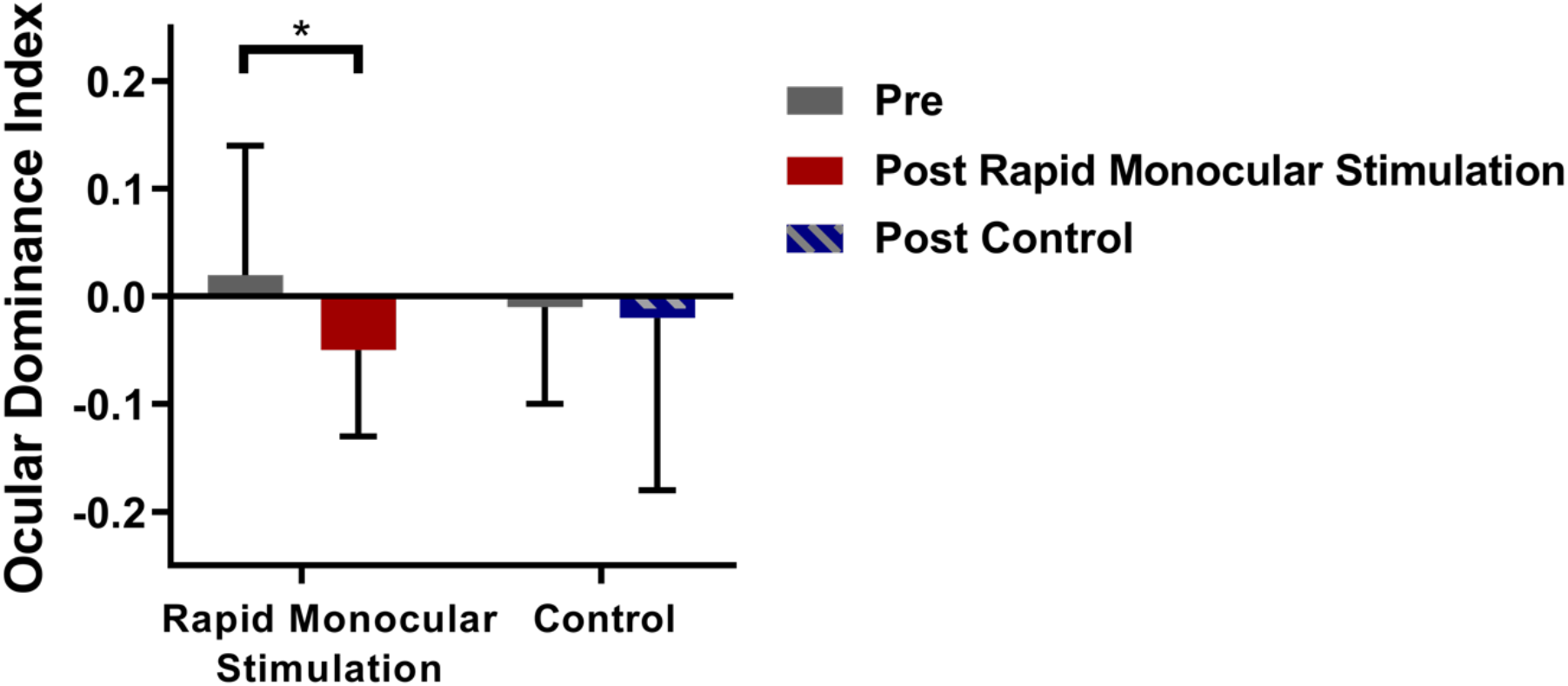
Median ocular dominance indices for the rapid visual stimulation and binocular control conditions in experiment 2. Positive values indicate increased dominance for the stimulated eye. Error bars = IQR.

### Experiment 3

Experiment 3 data were not normally distributed. For the subgroup from experiment 2 who also completed experiment 3, monocular adaptation did not alter ocular dominance (W = 30.0, p =0.838). Importantly, this subgroup did show a significant shift in ocular dominance towards the non-stimulated eye following rapid monocular stimulation, similar to the full cohort in experiment 2 (W = 66.0, p = 0.004; Figure 5). This subgroup also showed no effect of the binocular control condition (W = 32.0, p = 0.610).

**Figure 5:**
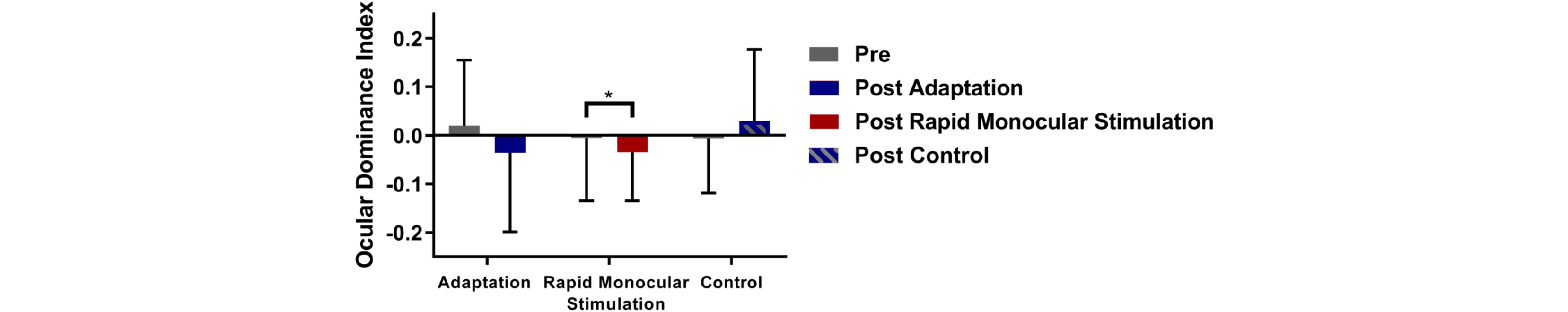
Median ocular dominance indices for Experiment 3. Positive values indicate increased dominance for the stimulated/adapted eye. Error bars = IQR.

## Discussion

The primary aim of this study was to assess whether rapid monocular stimulation of one eye would increase the dominance of that eye during binocular rivalry. Unexpectedly, we observed the opposite effect; rapid monocular stimulation *reduced* the relative dominance of the stimulated eye during binocular rivalry.

How might we explain this unexpected result? The simplest explanation is that rapid monocular stimulation caused retinal or cortical adaptation resulting in reduced dominance of the stimulated eye during binocular rivalry. Following previous work (Teyler *et al*., 2005), our rapid monocular stimulation protocol was designed to minimise adaptation effects by providing a period of eye closure directly after the rapid visual stimulation that was the same duration as the rapid visual stimulation itself (2 minutes). Generally, a period of adaptation lasts as long as the stimulation (Greenlee *et al*., 1991; see Başgöze *et al*., 2018 for an indepth review). However, it is still possible that adaptation played a role in our results. Therefore, we conducted a third experiment on a subset of participants from experiment 2 who were available and willing to complete further testing. This experiment revealed that simply adapting one eye to one of the gratings that made up the binocular rivalry stimulus did not alter ocular dominance. Together, the use of a period of eye closure within our rapid monocular stimulation protocol and the results of experiment 3 argue against adaptation as an explanation of our unexpected result.

An alternative explanation is that rapid visual stimulation of one eye may not have generated the expected LTP-like effects but rather a long-term depression-like effect (LTD). Although increased cortical excitability is the most commonly reported effect of visual stimulation (Teyler *et al*., 2005; Clapp *et al*., 2006; Kirk *et al*., 2010; de Gobbi Porto *et al*., 2015), decreased or inconsistent changes in cortical activity have also been reported. These include a reduced visual cortex BOLD response post-stimulation (Lahr *et al*., 2014) and reduced VEP amplitude in young adults post stimulation (Abuleil *et al*., 2019). The reason that some studies show LTP-like and others show LTD-like results is not clear; however, they do suggest that visual stimulation effects are inconsistent (Sanders *et al*., 2018). LTD-like changes following visual stimulation would be consistent with our observation of relatively reduced binocular rivalry dominance for the eye that received rapid monocular stimulation.

One additional possible explanation for decreased dominance following rapid monocular stimulation is suggested by recent studies that have explored the effect of short-monocular occlusion on binocular rivalry dominance. When one eye is occluded for a period of time, that eye has a relatively increased dominance during binocular rivalry once the occlusion is removed (Lunghi *et al*., 2011; Min *et al*., 2018). This effect does not require the deprived eye to be patched. Induced suppression of one eye or the presentation of lower contrast images to one eye for as little as 3 minutes also increases that eye’s binocular rivalry dominance (Kim *et al*., 2017). Other image degradation manipulations such as the presentation of pink noise (Bai *et al*., 2017) or spatial scrambling of one eye’s image also result in increased dominance of the deprived eye over the eye exposed to normal visual stimulation (Zhou *et al*., 2014; Ramamurthy & Blaser, 2018). The effects of short-term monocular occlusion also extend to participants with amblyopia, a disorder characterised by chronic perceptual dominance of the fellow eye over the amblyopic eye (Li *et al*., 2011). Occlusion of the amblyopic eye strengthens the contribution of that eye to binocular vision once the occlusion is removed (Lunghi *et al*., 2011, 2016; Zhou *et al*., 2013; Chadnova *et al*., 2017). Possible mechanisms underlying the ocular dominance shift induced by short-term monocular occlusion include a change in neural interocular gain control resulting from a large imbalance in the input from each eye to cortical processing (Lunghi *et al*., 2011; Zhou *et al*., 2013). This change is associated with reduced visual cortex GABA concentration (Lunghi *et al*., 2015) and may involve both feedforward and feedback pathways (Ramamurthy & Blaser, 2018).

We postulate that the strengthening of the cortical response to the stimulated eye generated by our monocular rapid stimulation protocol activated the same homeostatic mechanisms that underpin short-term monocular occlusion effects. In other words, the reduced binocular rivalry dominance of the stimulated eye was not a direct effect of the rapid monocular stimulation but was caused by the relative deprivation of the non-stimulated eye. This raises the exciting possibility that rapid monocular stimulation can be used to rapidly induce eye dominance shifts. Potential applications of this technique include the manipulation of ocular dominance in amblyopia. We are currently conducting studies that address this possibility.

## Data Availability

Data is available when requested

## References

Abuleil, D., McCulloch, D.L., & Thompson, B. (2019) Older adults exhibit greater visual cortex inhibition and reduced visual cortex plasticity compared to younger adults. Front. Neurosci., 13, 1–7.

Bai, J., Dong, X., He, S., & Bao, M. (2017) Monocular deprivation of Fourier phase information boosts the deprived eye’s dominance during interocular competition but not interocular phase combination. Neuroscience, 352, 122–130.

Barnes, G.R., Hess, R.F., Dumoulin, S.O., Achtman, R.L., & Pike, G.B. (2001) The cortical deficit in humans with strabismic amblyopia. J. Physiol., 533.1, 281–297.

Başgöze, Z., Mackey, A.P., & Cooper, E.A.(2018) Plasticity and adaptation in adult binocular vision. Curr. Biol., 28, R1406–R1413.

Beste, C., Wascher, E., Güntürkün, O., & Dinse, H.R. (2011) Improvement and impairment of visually guided behavior through LTP-and LTD-like exposure-based visual learning. Curr. Biol., 21, 876–882.

Blakemore, C. & Campbell, F.W. (1969) On the existence of neurones in the human visual system selectively sensitive to the orientation and size of retinal images. J. Physiol.,203, 237–260.

Bliss, T.V.P. & Collingridge, G.L. (1993) A synaptic model of memory: long-term potentiation in the hippocampus. Nature, 361, 31–39.

Bliss, T.V.P. & Lømo, T. (1973) Long-lasting potentiation of synaptic transmission in the dentate area of the unanaestetized rabbit following stimulation of the perforant path. J. Physiol., 232, 357–374.

Bröcher, S., Artola, A., & Singer, W. (1992) Agonists of cholinergic and noradrenergic receptors facilitate synergistically the induction of long-term potentiation in slices of rat visual cortex. Brain Res., 573, 27–36.

Chadnova, E., Reynaud, A., Clavagnier, S., & Hess, R.F. (2017) Short-term monocular occlusion produces changes in ocular dominance by a reciprocal modulation of interocular inhibition. Sci. Rep., 7, 2–7.

Clapp, W., Zaehle, T., Lutz, K., Marcar, V., Kirk, I.J., Hamm, J.P., Teyler, T.J., Corballis, M.C., & Jancke, L. (2005) Effects of long-term potentiation in the human visual cortex: a functional magnetic resonance imaging study. Neuroreport, 16, 1977–1980.

Clapp, W.C., Eckert, M.J., Teyler, T.J., & Abraham, W.C. (2006) Rapid visual stimulation induces N-methyl-D-aspartate receptor-dependent sensory long-term potentiation in the rat cortex. Neuroreport, 17, 511–515.

Clapp, W.C., Hamm, J.P., Kirk, I.J., & Teyler, T.J. (2012) Translating long-term potentiation from animals to humans: A novel method for noninvasive assessment of cortical plasticity. Biol. Psychiatry, 71, 496–502.

Clavagnier, S., Thompson, B., & Hess, R.F. (2013) Long lasting effects of daily theta burst rTMS sessions in the human amblyopic cortex. Brain Stimul., 6, 860–867.

de Gobbi Porto, F.H., Fox, A.M., Tusch, E.S., Sorond, F., Mohammed, A.H., & Daffner, K.R. (2015) In vivo evidence for neuroplasticity in older adults. Brain Res. Bull., 114, 56–61.

Eckert, M.J., Guévremont, D., Williams, J.M., & Abraham, W.C. (2013) Rapid visual stimulation increases extrasynaptic glutamate receptor expression but not visual-evoked potentials in the adult rat primary visual cortex. Eur. J.Neurosci., 37, 400–406.

Fahle, M. (1982) Binocular rivalry: Suppression depends on orientation and spatial frequency. Vision Res., 22, 787–800.

Frenkel, M.Y., Sawtell, N.B., Diogo, A.C.M., Yoon, B., Neve, R.L., & Bear, M.F. (2006) Instructive effect of visual experience in mouse visual cortex. Neuron, 51, 339–349.

Greenlee, M.W., Georgeson, M.A., Magnussen, S., & Harris, J.P. (1991) The time course of adaptation to spatial contrast. Vision Res., 31, 223–236.

Hayashi, Y., Shi, S.H., Esteban, J. a, Piccini, a, Poncer, J.C., & Malinow, R. (2000) Driving AMPA receptors into synapses by LTP and CaMKII: requirement for GluR1 and PDZ domain interaction. Science, 287, 2262–2267.

Heynen, A.J. & Bear, M.F. (2001) Long-term potentiation of thalamocortical transmission in the adult visual cortex in vivo. J. Neurosci., 21,9801–9813.

Holmes, D.J., Hancock, S., & Andrews, T.J. (2006) Independent binocular integration for form and colour. Vision Res., 26,665–677.

Hong, S.W. & Shevell, S.K. (2008) The influence of chromatic context on binocular color rivalry: Perception and neural representation. Vision Res., 48,1074–1083.

Hoogendam, J.M., Ramakers, G.M.J., & Di Lazzaro, V. (2010) Physiology of repetitive transcranial magnetic stimulation of the human brain. Brain Stimul., 3, 95–118.

Kang, M.-S.S. (2009) Size matters: a study of binocular rivalry dynamics. J. Vis., 9, 1–11.

Kim, H.W., Kim, C.Y., & Blake, R. (2017) Monocular perceptual deprivation from interocular suppression temporarily imbalances ocular dominance. Curr. Biol., 27,884–889.

Kirk, I.J., McNair, N.a., Hamm, J.P., Clapp, W.C., Mathalon, D.H., Cavus, I., & Teyler, T.J. (2010) Long-term potentiation (LTP) of human sensory-evoked potentials. Wiley Interdiscip. Rev. Cogn. Sci., 1,766–773.

Lahr, J., Peter, J., Bach, M., Mader, I., Nissen, C., Normann, C., Kaller, C.P., & Klöppel, S. (2014) Heterogeneity of stimulus-specific response modification-an fMRI study on neuroplasticity. Front. Hum. Neurosci., 8,695.

Li, J., Thompson, B., Lam, C.S.Y., Deng, D., Chan, L.Y.L., Maehara, G., Woo, G.C., Yu, M., & Hess, R.F. (2011) The role of suppression in amblyopia. Invest. Ophthalmol. Vis. Sci., 52,4169–4176.

Lunghi, C., Berchicci, M., Morrone, M.C., & Di Russo, F. (2015) Short-term monocular deprivation alters early components of visual evoked potentials. J. Physiol., 593,4361–4372.

Lunghi, C., Burr, D.C., & Morrone, C. (2011) Brief periods of monocular deprivation disrupt ocular balance in human adult visual cortex. Curr. Biol.,21, R538–R539.

Lunghi, C., Morrone, M.C., Secci, J., & Caputo, R. (2016) Binocular rivalry measured 2 hours after occlusion therapy predicts the recovery rate of the amblyopic eye in anisometropic children. Investig. Ophthalmol. Vis. Sci., 57,1537–1546.

Magnussen, S. & Greenlee, M.W. (1985) Marathon adaptation to spatial contrast: Saturation in sight. Vision Res., 25,1409–1411.

Malenka, R.C. & Nicoll, R.A. (1999) Long-term potentiation – A decade of progress? Sci. Mag., 285,1870–1874.

Min, S.H., Baldwin, A.S., Reynaud, A., & Hess, R.F. (2018) The shift in ocular dominance from short-term monocular deprivation exhibits no dependence on duration of deprivation. Sci. Rep., 8,1–9.

Normann, C., Schmitz, D., Fürmaier, A., Döing, C., & Bach, M. (2007) Long-term plasticity of visually evoked potentials in humans is altered in major depression. Biol. Psychiatry, 62,373–380.

Ramamurthy, M. & Blaser, E. (2018) Assessing the kaleidoscope of monocular deprivation effects. J. Vis., 18,14.

Ross, R.M., McNair, N.a, Fairhall, S.L., Clapp, W.C., Hamm, J.P., Teyler, T.J., & Kirk, I.J. (2008) Induction of orientation-specific LTP-like changes in human visual evoked potentials by rapid sensory stimulation. Brain Res. Bull., 76,97–101.

Sanders, P.J., Thompson, B., Corballis, P.M., Maslin, M., & Searchfield, G.D. (2018) A review of plasticity induced by auditory and visual tetanic stimulation in humans. Eur.J.Neurosci., 2084–2097.

Stalmeier, P.F.M. & de Weert, C.M.M. (1988) Binocular rivalry with chromatic contours. Percept. Psychophys., 44,456–462.

Teyler, T.J. & DiScenna, P. (1987) Long-term potentiation. Annu. Rev. Neurosci., 10,131–161.

Teyler, T.J., Hamm, J.P., Clapp, W.C., Johnson, B.W., Corballis, M.C., & Kirk, I.J. (2005) Long-term potentiation of human visual evoked responses. Eur.J.Neurosci., 21,2045–2050.

Thompson, B., Mansouri, B., Koski, L., & Hess, R.F. (2008) Brain Plasticity in the Adult: Modulation of Function in Amblyopia with rTMS. Curr. Biol., 18,1067–1071.

Tuna, A.R., Pinto, N., Brardo, F.M., Fernandes, A., Nunes, A.F., & Pato, M.V. (2020) Transcranial magnetic stimulation in adults with amblyopia. J.Neuro-Ophthalmology, 40,185–192.

Vassilev, A., Stomonyakov, V., & Manahilov, V. (1994) Spatial-frequency specific contrast gain and flicker masking of human transient VEP. Vision Res.,34,863–872.

Wilson, H.R., Blake, R., & Lee, S.H. (2001) Dynamics of travelling waves in visual perception. Nature, 412,907–910.

Zhou, J., Clavagnier, S., & Hess, R.F. (2013) Short-term monocular deprivation strengthens the patched eye’s contribution to binocular combination. J. Vis., 13,1–10.

Zhou, J., Reynaud, A., & Hess, R.F. (2014) Real-time modulation of perceptual eye dominance in humans. Proc.R.Soc. B Biol. Sci.,281,1–6.

